# Early life phthalate exposure impacts gray matter and white matter volume in infants and young children

**DOI:** 10.1101/2025.02.05.25321734

**Authors:** Emily J. Werder, Kun Lu, Chih-Wei Liu, Jake E. Thistle, Julia E. Rager, Gang Li, Zhengwang Wu, Tengfei Li, Li Wang, Dale P. Sandler, John H. Gilmore, Joseph Piven, Hongtu Zhu, Weili Lin, Stephanie M. Engel

## Abstract

**Objective:** Prenatal phthalate exposure is associated with adverse neurodevelopmental outcomes, yet data on impacts of early life exposure remains limited. We investigated phthalate and replacement plasticizer exposures from 2 weeks to 7 years of age in relation to brain anatomical attributes, using serial structural magnetic resonance imaging (sMRI).

**Material and Methods:** Children were enrolled after birth into the UNC Baby Connectome Project, a longitudinal neuroimaging study. Urine samples (n=406) were collected at each visit and analyzed for 17 phthalate and replacement plasticizer metabolites. Among 157 children contributing 369 sMRIs, we calculated metabolite-specific average exposures across each individual’s urine samples and used linear mixed models to estimate longitudinal associations of log transformed, specific gravity-adjusted average metabolite concentrations with gray (GMV) and white matter (WMV), and cortical volume (CV), thickness (CT), and surface area (CSA). We examined sex-specific differences in these associations.

**Results:** Higher average metabolite concentration was associated with lower GMV (MCPP: (−1.73 cm^3^, 95% CI: −3.36, −0.10) and higher WMV (∑DEHP: 2.28 cm^3^, 95% CI: 0.08, 4.48). Among boys (n=72, 140 sMRIs), MEP (−2.97 cm^3^, 95% CI: −5.85, −0.09) and MiBP (−2.40 cm^3^, 95% CI: −4.64, −0.15) were also associated with lower GMV. Among girls (n=85, 229 MRIs), higher ∑DINCH exposure was associated with higher WMV (2.27 cm^3^, 95% CI: 0.29, 4.25). We observed significant sex interactions for ∑DEHP with GMV (p-interaction=0.03) and ∑DINCH with WMV (p-interaction=0.001).

**Conclusion:** Early life phthalate/plasticizer exposure may differentially impact various brain region volumes in early childhood, with potential downstream consequences on functional development.

## Introduction

The first years of life are a critical period of accelerated brain growth that lay the foundation for lifelong neurodevelopmental trajectories of cognition and behavior, as well as risk for neurodevelopmental disorders. This period of accelerated gray matter growth, rapid myelination of white matter, and maturation of structural and functional networks^1–3^ coincides with the emergence of behavioral and cognitive abilities^4^. Because brain development occurs via a precisely synchronized sequence of coordinated events, exposure to environmental toxicants at critical stages may disrupt essential processes, initiating a deleterious cascade that fundamentally alters neurobehavioral trajectories.

Children are exposed to a myriad of xenobiotic neurotoxicants^5^ in air, water, and dietary sources^6–8^. The infant brain may be especially vulnerable to toxicants because fragile cerebral vessels in the neurovascular unit impact permeability of the blood brain barrier,^9^ and immature toxicant metabolic systems constrain metabolism and excretion of toxicants^10,11^. Infants no longer have the protection that was provided by the placenta during fetal development^9^, which may render them more vulnerable to toxicant exposures than the fetus. Infants and children are exposed to chemicals through hand-to-mouth behaviors, increased contact with the ground, breastfeeding, formula consumption, sustained use of baby products, and their diet.

Phthalates are a class of synthetic chemicals that are widely used as solvents and plasticizers in commercial and consumer products, such as medical devices, personal care products and cosmetics, food packaging, and building materials. Human exposure to phthalates is ubiquitous, occurring through dermal absorption, inhalation, and ingestion. Diet is a principal exposure pathway^12^. Due to mounting evidence that di(2-ethylhexyl) phthalate (DEHP) and dibutyl phthalate (DBP) are harmful to human health, their use has been restricted in children’s toys and other childcare products in the United States. Commonly used phthalate replacement plasticizers, including di(isononyl)cyclohexane-1,2-dicarboxylate (DINCH) and di-2-ethylhexyl terephthalate (DEHTP)^13,14^, are increasingly found in population based sampling of the United States^15–17^. In addition, we previously reported widespread detection of urinary metabolites of phthalates and replacements plasticizers in infancy and early childhood in the University of North Carolina (UNC) Baby Connectome Project (BCP)^18^. Although NHANES human biomonitoring data in the general US population includes only children ages 3 and older, available data indicate that younger age groups typically have higher concentrations of phthalate metabolites^19,20^. Indeed, in our own study, we observed age-related differences in exposure wherein phthalate levels were often highest in the first year of life and declined with age^18^.

Robust evidence demonstrates that prenatal phthalate exposure elicits adverse neurobehavioral, cognitive, and developmental impacts^21–24^, yet the structural and functional substrates underlying these relationships remain understudied. The few neuroimaging studies which have evaluated the role of phthalate exposure in neurodevelopment assess heterogeneous endpoints at varying ages^25^. England-Mason et al. evaluated prenatal phthalates and white matter microstructure at 3-5 years of age in the Alberta Pregnancy Outcomes and Nutrition (APrON) study (n=76)^26^, reporting associations between high molecular weight phthalates and increased mean diffusivity of white matter tracts. In the Generation R study (n=775), prenatal phthalates were associated with global brain volumes at age 10, with more prominent effects among girls^27^. In a study of 49 teenagers, prenatal phthalates were associated with reduced regional brain volumes and altered white matter integrity at 13-16^28^. The only existing study of postnatal phthalate exposure and neuroimaging, a cross-sectional analysis of 8–11-year-olds diagnosed with attention deficit hyperactivity disorder (n=115), reported that metabolites of di(2-ethylhexyl) phthalate (DEHP) were negatively correlated with cortical thickness in the right middle and superior temporal gyri^29^.

The nascent body of research investigating the links between phthalate exposure and neuroimaging biomarkers is largely restricted to prenatal exposures in relation to MRIs acquired later in adolescence. Studies linking prospective postnatal exposure, longitudinal scans, and imaging during sensitive periods of early development are needed to better understand the impacts of phthalate exposure on neurodevelopment during this critical window of development^30^. To address these gaps, we examined early life phthalate and replacement plasticizer exposure in relation to global measures of structural brain growth from two weeks to seven years of age in the UNC BCP. Based on previous evidence that neurodevelopmental effects of phthalate exposure are sexually dimorphic, we additionally evaluated differences in associations between early life phthalate exposure and global brain measures by child sex.

## Methods

### Study population

The BCP is a longitudinal study that aims to characterize normative brain and behavioral development in typically developing infants across the first 5 years of life^31^. Eligible participants were born between 37-42 weeks’ gestation, with birth weight appropriate for gestational age, absence of pregnancy complications, and parents reported no medical or genetic conditions related to neurodevelopment. The BCP enrolled participants between birth and age five at two sites, the University of Minnesota and the University of North Carolina – Chapel Hill (UNC), between 2017 and 2020. To limit participant attrition, the BCP employed an accelerated cohort design, with infants and children entering observation in a staggered pattern and followed forward with longitudinal scans spanning different periods of early life. This dense longitudinal sampling within developmental periods maximizes coverage across a wider age range while relying on a shorter follow-up period (Supplemental Figure 1).

**Figure 1.**
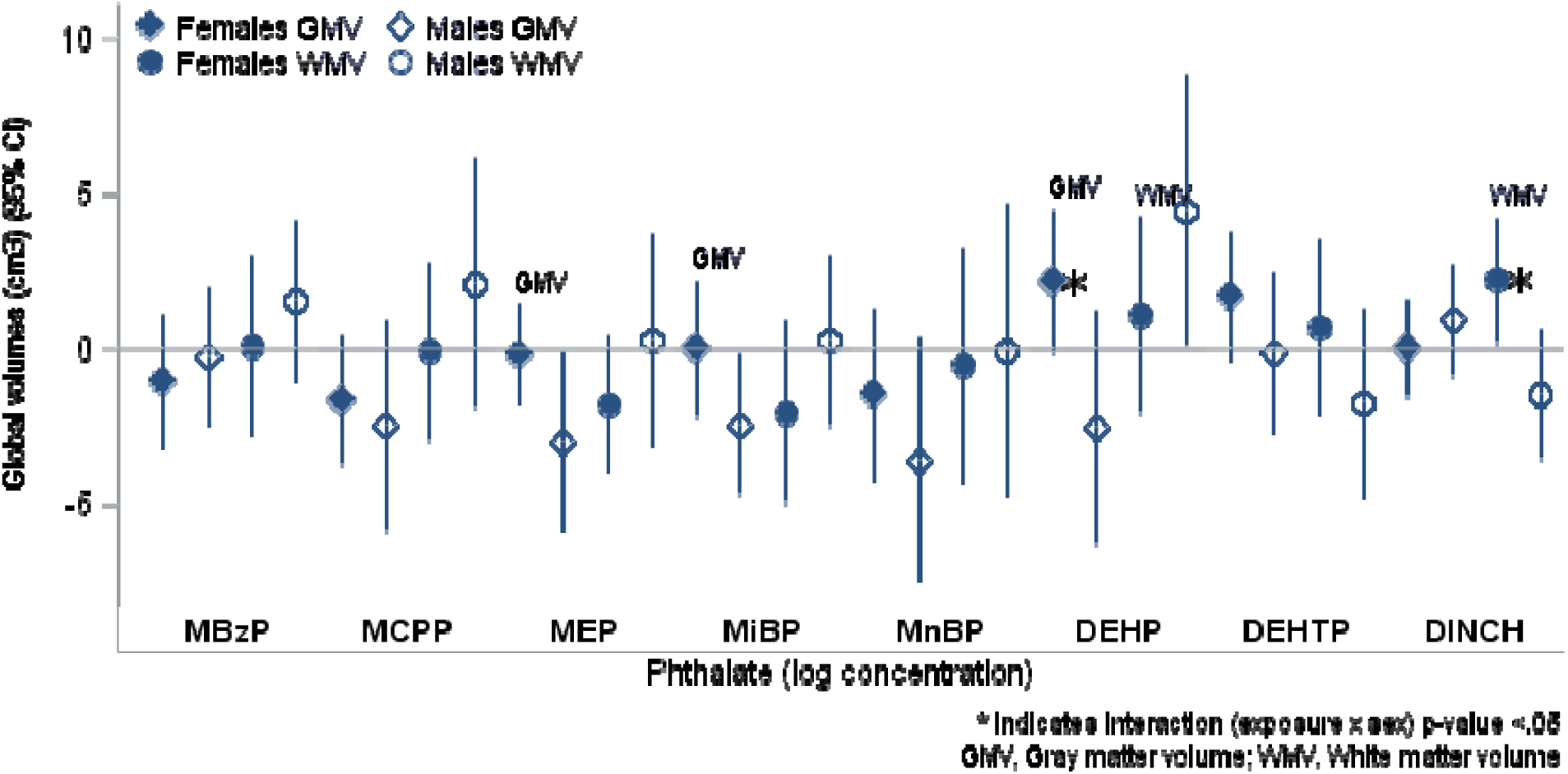
Associations of total average phthalate exposure and global brain volumes by sex in the UNC BCP (n=229 females and 140 males). GMV, gray matter volume (cm^3^); WMV, white matter volume (cm^3^); CI, confidence interval; MBzP, monobenzyl phthalate; MCPP, mono-3carboxypropyl phthalate; MEP, monoethyl phthalate; MiBP, monoisobutyl phthalate; MnBP, monobutyl phthalate; DEHP, di(2-ethylhexyl) phthalate; DEHTP, di-2-ethylhexyl terephthalate; DINCH, di(isononyl)cyclohexane-1,2-dicarboxylate; effect estimates are per log-unit increase in SG-adjusted average urinary phthalate concentrations (log(ng/mL) for metabolites and log(µmol/L) for molar sums); models are stratified by child’s sex and adjusted for maternal age at delivery, child’s age (days) on the date of the MRI, and intracranial volume; interaction p-value is for product term between phthalate exposure and child’s sex in augmented product term model.

The UNC BCP site (n = 238) additionally attempted to collect a urine sample at every participant visit. The present study is limited to UNC BCP participants who provided at least one urine sample (n=187) for exposure ascertainment, completed at least one MRI (n=159), and had known covariate information, for an analytic sample of 157 participants contributing 406 urines and 369 scans.

### Urine sample collection

Urine samples were collected at each attempted imaging appointment. For infants and young children wearing diapers, research assistants provided parents with a fresh diaper (Bambo Nature Premium) and Waterwipes to remove residual lotion or creams prior to placing 4–5 hospital-grade cotton balls in the diaper at the beginning of the visit. After the visit, cotton balls were collected in a sterile, phthalate-free container and stored at 4 °C. For toilet trained children, a sterile phthalate-free toilet hat was placed in the toilet to catch urine at any time during the visit. Once collected, samples were sent to the UNC BioSpecimen Processing Facility (median processing time ~15 h). Urine was extracted from cotton balls using a phthalate-free syringe and stored securely. Samples were aliquoted and stored at −80 °C within 24 h of processing.

### Phthalate exposure measurement

The detailed protocol for urine sample preparation and mass spectrometry analysis have been described previously^32^. Briefly, concentrations were quantified using highly sensitive triple quadrupole mass spectrometry with ultra-high performance liquid chromatography for 17 metabolites of phthalates and replacement plasticizers: monobutyl phthalate (MnBP), mono-3carboxypropyl phthalate (MCPP), monoisobutyl phthalate (MiBP), monoethyl phthalate (MEP), monobenzyl phthalate (MBzP), mono-2ethylhexyl phthalate (MEHP), mono-2-ethyl-5-hydroxyhexyl phthalate (MEHHP), mono-2-ethyl-5-oxohexyl phthalate (MEOHP), mono-2-ethyl5-carboxypentyl phthalate (MECPP), mono-2-carboxymethylhexyl phthalate (MCMHP), monooxononyl phthalate (MONP), monoisononyl phthalate (MCOP), monocarboxyisononyl phthalate (MCNP), cyclohexane-1,2-dicarboxylic acid, monohydroxy isononyl ester (MHNCH), cyclohexane-1,2-dicarboxylic acid, monocarboxy isooctyl ester (MCOCH), mono-2-ethyl-5-hydrohexyl terephthalate (MEHHTP), and mono-2-ethyl-5-carboxypentyl terephthalate (MECPTP). All metabolite concentrations were adjusted for specific gravity (SG)^18^, an appropriate approach to account for urinary dilution in infants and young children^33,34^. Concentrations below the limit of detection (LOD) were imputed using the LOD/√2. We calculated molar sums (μmol/L) of metabolite concentrations by summing SG-adjusted and LOD/√2-imputed metabolite concentrations after dividing by the molecular weight, for three compounds: DEHP (∑DEHP = MEHP + MECPP + MEHHP + MEOHP + MCMHP); DINCH (∑DINCH = MCOCH + MHNCH); and DEHTP (∑DEHTP = MECPTP + MEHHTP). Association analyses were completed for eight distinct exposures: all five metabolites (MnBP, MCPP, MiBP, MEP, MBzP) and three molar sums (DEHP, DINCH, DEHTP).

### Neuroimaging acquisition and processing

The BCP imaging protocol^31^, which was based on the imaging protocol developed by the Human Connectome Project^35^, consists of sMRI T_1_-weighted (T_1_w) and T_2_-weighted (T_2_w), resting-state functional MRI (rsfMRI), and diffusion MRI (dMRI). Images were acquired on 3T Siemens Prisma MRI scanners using a Siemens 32 channel head coil at the UNC Biomedical Research Imaging Center (BRIC). If considerable motion was observed at the time of scanning, study personnel attempted reacquisition if possible.

Structural MR images were processed by iBEAT V2.0 pipeline (http://www.ibeat.cloud)^36^. sMRI data were assessed visually for excessive motion, insufficient coverage, and ghosting. The sMRI T_1_w and T_2_w images provide information regarding structural brain development. Outcomes of interest included global volumes (cm^3^): gray matter (GMV), white matter (WMV), and cortical gray matter (CV); cortical thickness (CT, mm), and cortical surface area (CSA, cm^2^). We used intracranial volume (cm^3^) as an adjustment factor for scanning effects.

### Statistical analysis

For phthalate and replacement plasticizer exposures, we log transformed concentrations and then averaged across all samples provided by an individual. Thus, we assigned each participant eight exposure metrics, each corresponding to the average concentration of a specific metabolite or molar sum. We additionally calculated cumulative average exposures for each exposure metabolite or molar sum at each MRI, averaging concentrations from all of an individual’s samples until the date of the index MRI (but excluding those provided after the scan date). Because phthalates are rapidly metabolized and have short half-lives^37^, we prioritized the *total* average concentrations (as opposed to *cumulative* average concentrations) to leverage all available exposure information, better represent typical early life exposure, and maximize sample size and statistical power in primary analyses. Cumulative average exposures were analyzed in sensitivity analyses (n=284).

To account for repeated scans, and the correlation introduced with multiple observations per participant, we used linear mixed models with temporal power covariance to estimate longitudinal associations between average phthalate/plasticizer exposure and each of the global sMRI region outcomes. All models were adjusted for total intracranial volume, child’s sex assigned at birth, maternal age at delivery (continuous years), and child’s age on the date of the MRI (days). Due to the strong age-dependence of brain growth during early childhood, we adjusted for child’s age using natural cubic splines with knots at the 10^th^ (115 days), 25^th^ (335 days), 75^th^ (1,198 days), and 90^th^ (1,713 days) percentile. For white matter volume, we included an additional knot at the 5^th^ percentile (61 days) to improve model fit for this outcome. Knot selection and model specification were guided by goodness-of-fit tests and graphical interpretation of residuals. We modeled each exposure-outcome combination separately, reporting effect estimates as the amount of change in MRI feature (β coefficient) associated with a log-unit increase in exposure, and corresponding 95% confidence intervals (CIs). To account for multiple hypothesis testing, we report false discovery rate (FDR) corrected p-values for 40 tests (8 exposures and 5 outcomes)^38^.

Because phthalates are endocrine disrupting chemicals^39^ and early life brain growth trajectories are sex-dependent^40^, we estimated sex-specific associations in stratified models restricted to males (n=72 participants, 140 observations) or females (n=85 participants, 229 observations) with the same covariate adjustment set as the overall analysis. We evaluated sex differences in the overall sample using the augmented product term approach^41^, reporting the p-value associated with the two-sided Wald test statistic (alpha=0.05) for the product term between exposure and sex.

Sensitivity analyses evaluated time-varying cumulative average exposures and exposure-age interactions dichotomized at 12-,18-, and 24-months. All statistical analyses were conducted in SAS version 9.4 (Cary, NC, USA).

## Results

The sex distribution in the study sample was approximately balanced (54% female), though females contributed slightly more observations (n=229, 62% of scans) (Table 1). The youngest and oldest ages at MRI were at 6 days and 2,567 days (~7 years old), respectively. The majority of scans (61%) were obtained before age 2 years (Table 1). Participants contributed a median of 2 MRIs (maximum=8) and 2 urine samples (maximum=7). Detection rates for individual metabolites ranged from 83% (MEP) to 100% (MBzP, MnBP), and from 66% (DINCH) to 100% for molar sums (data not shown).

**Table 1.**
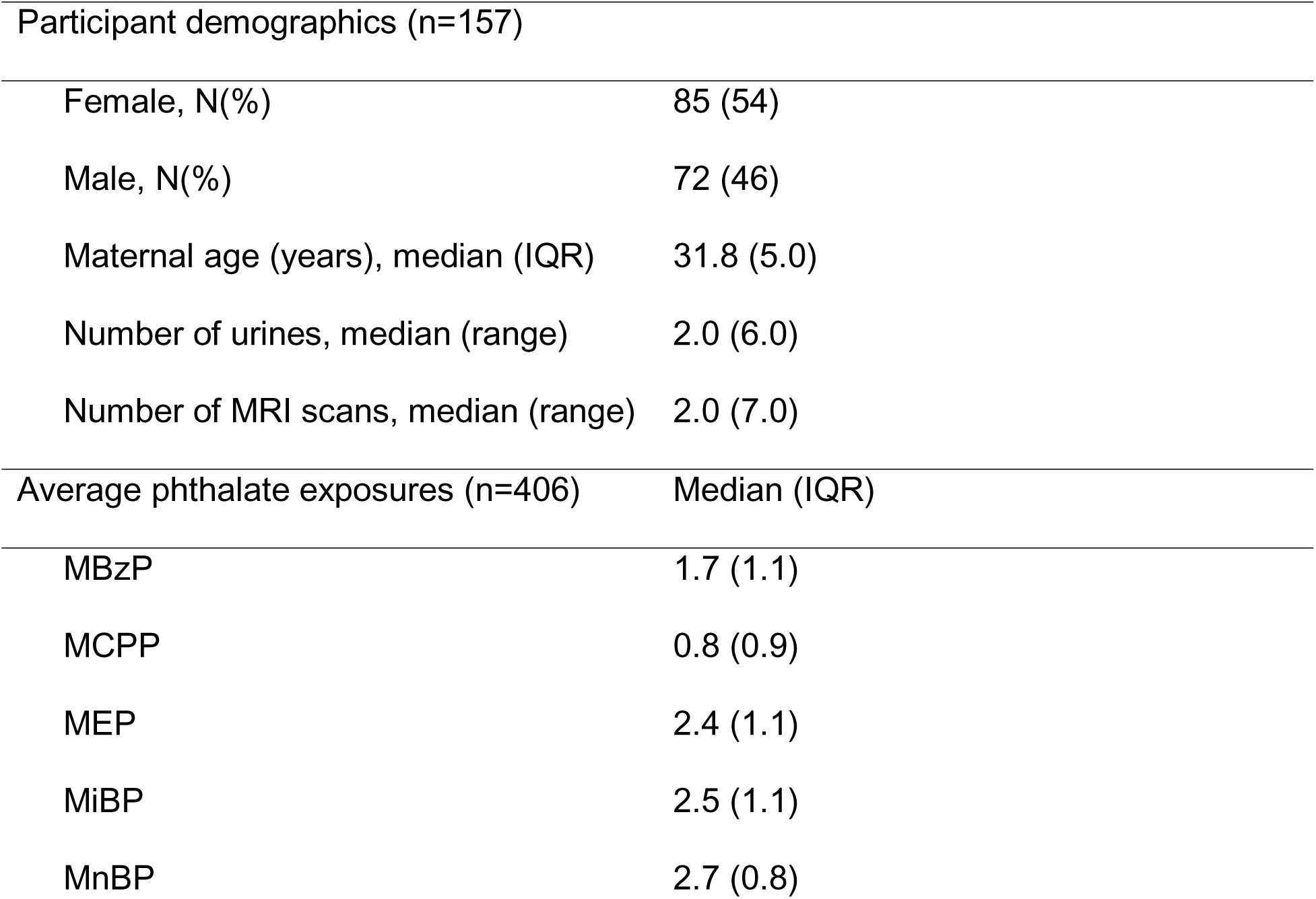
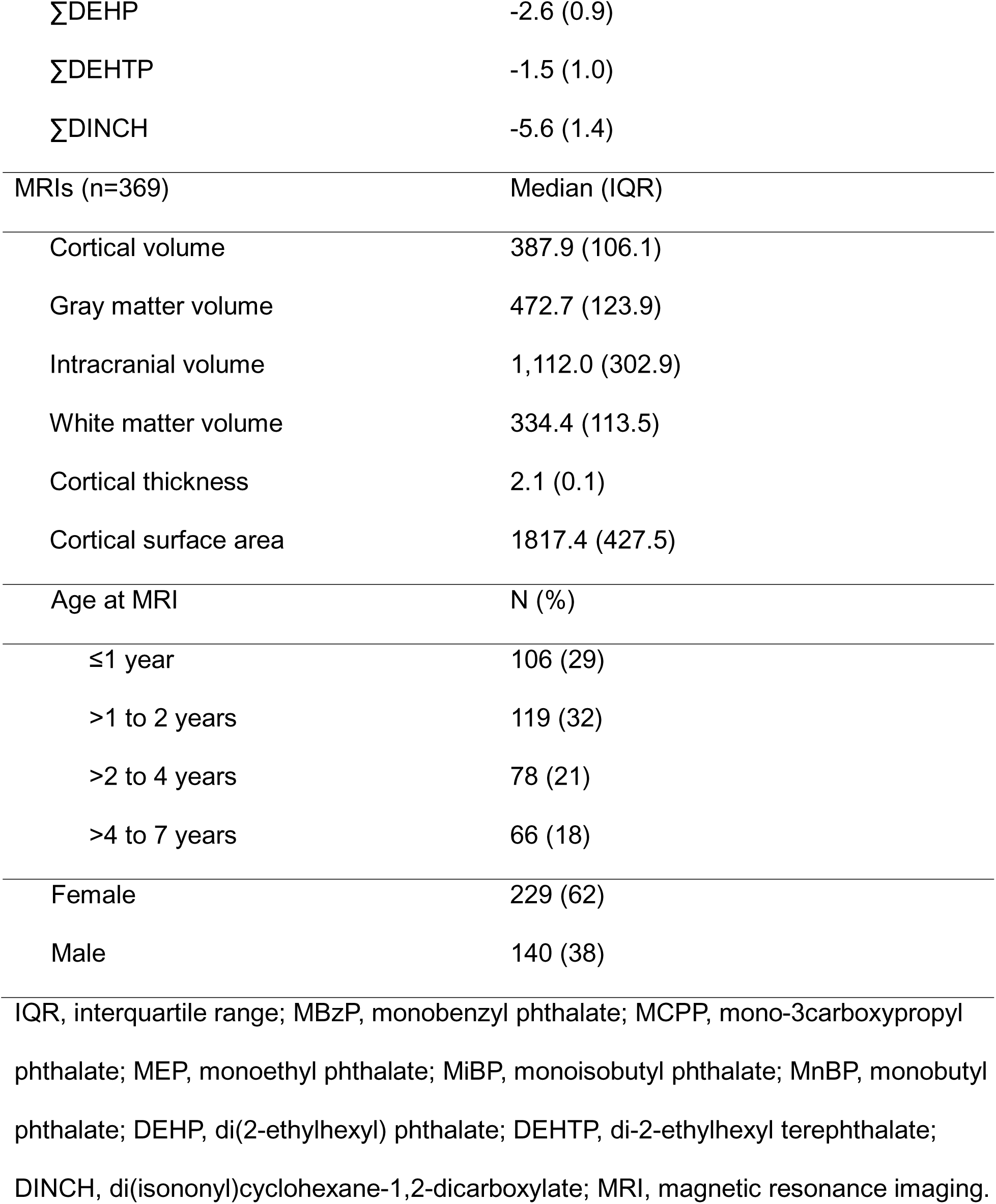
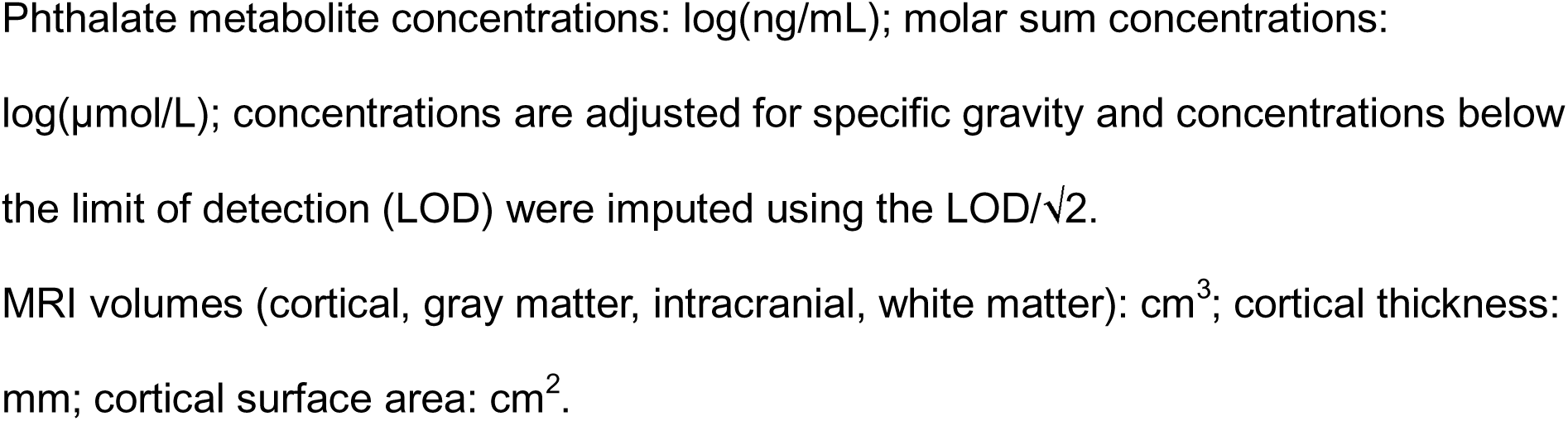
UNC Baby Connectome Project cohort Participant demographics (n=157)

In the overall sample, MCPP was associated with lower GMV (−1.7 cm^3^, 95% CI: −3.4, −0.1), whereas ∑DEHP was associated with higher WMV (2.3 cm^3^, 95% CI: 0.1, 4.5) (Table 2). Although not statistically significant, MnBP was suggestively associated with lower GMV (−2.0 cm^3^, 95% CI: −4.1, 0.05). We did not observe any significant associations with CV (Table 3), though patterns were similar between GMV and CV. Exposure associations with CV were generally slightly attenuated and less precise compared to corresponding estimates for GMV. MiBP was associated with lower cortical thickness (−0.01 mm, 95% CI: −0.02, −0.004). While no exposures were significantly associated with cortical surface area, ∑DEHP was associated with a 10.4 cm^2^ increase in CSA (95% CI: −0.02, 20.9).

**Table 2.**
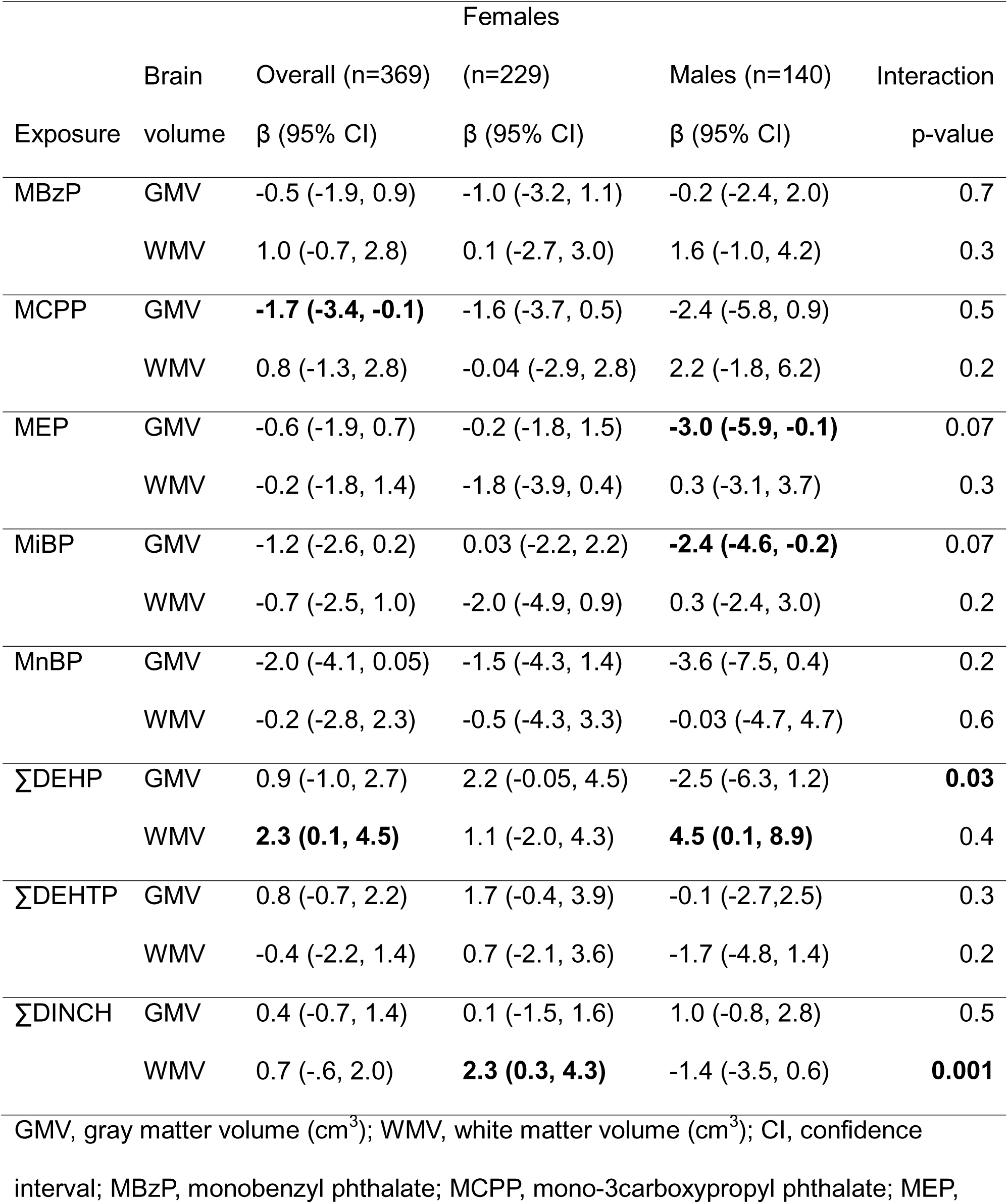
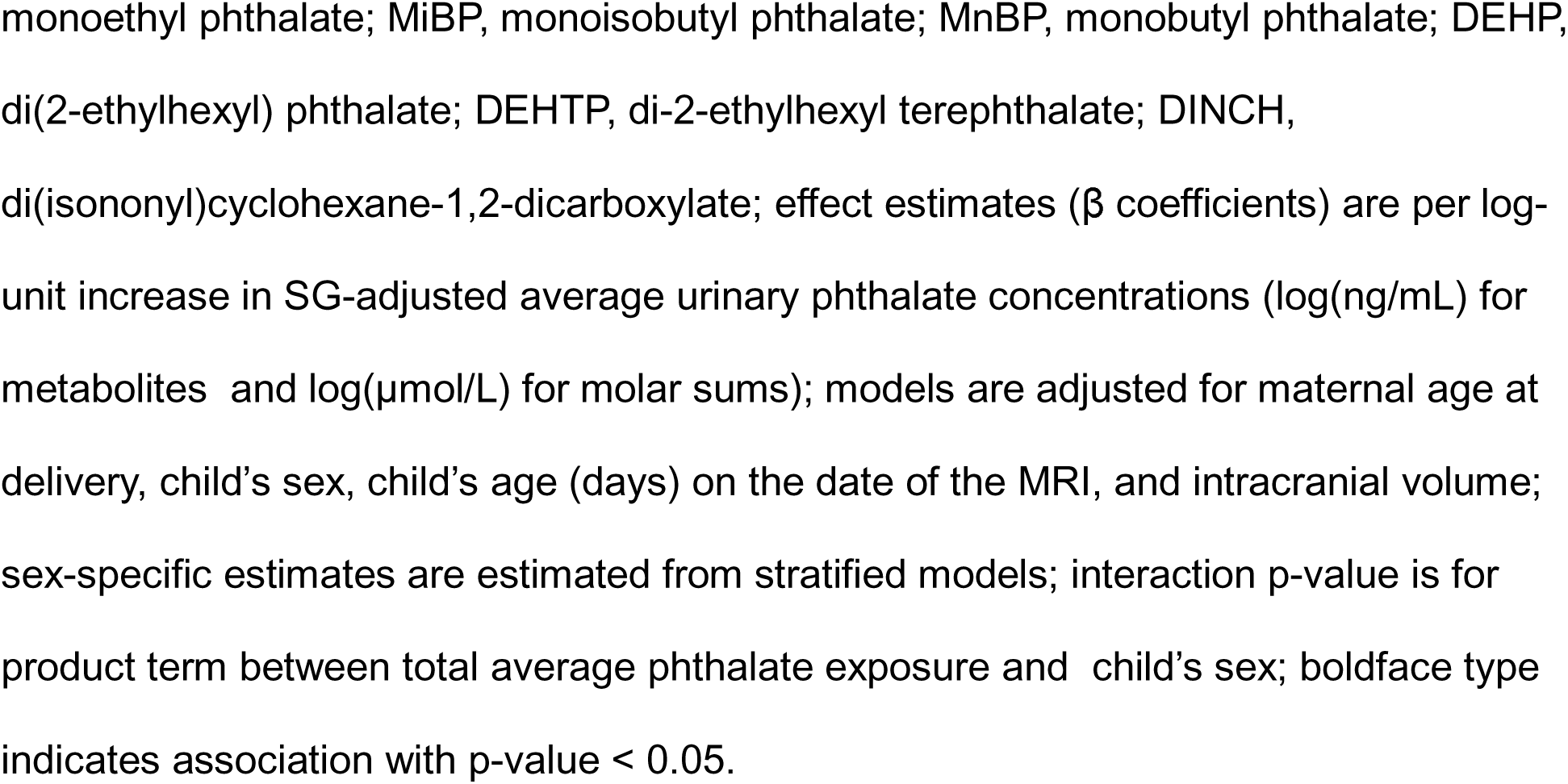
Associations of total average phthalate exposures with global gray and white matter volumes in the UNC BCP (n=369).

**Table 3.**
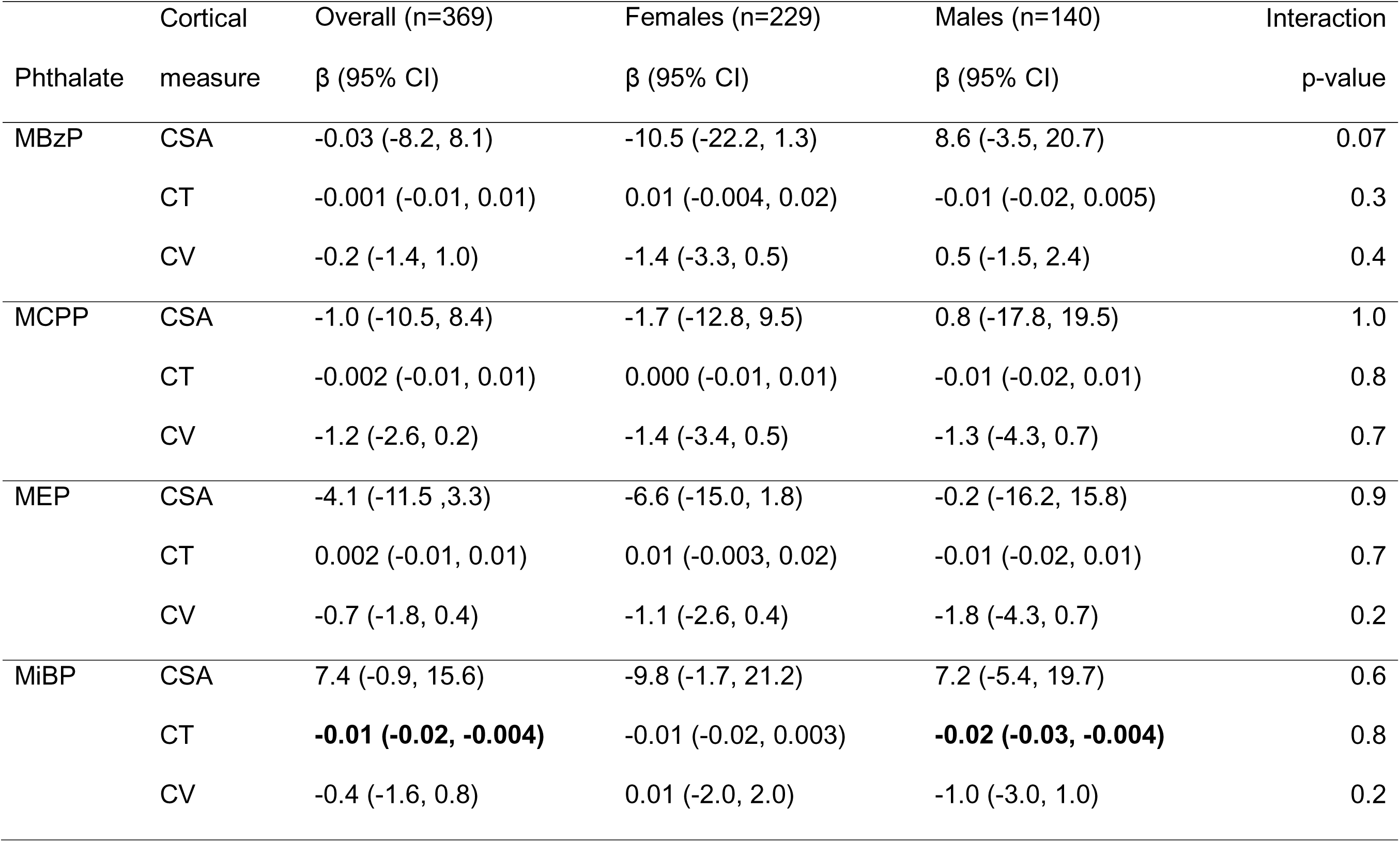
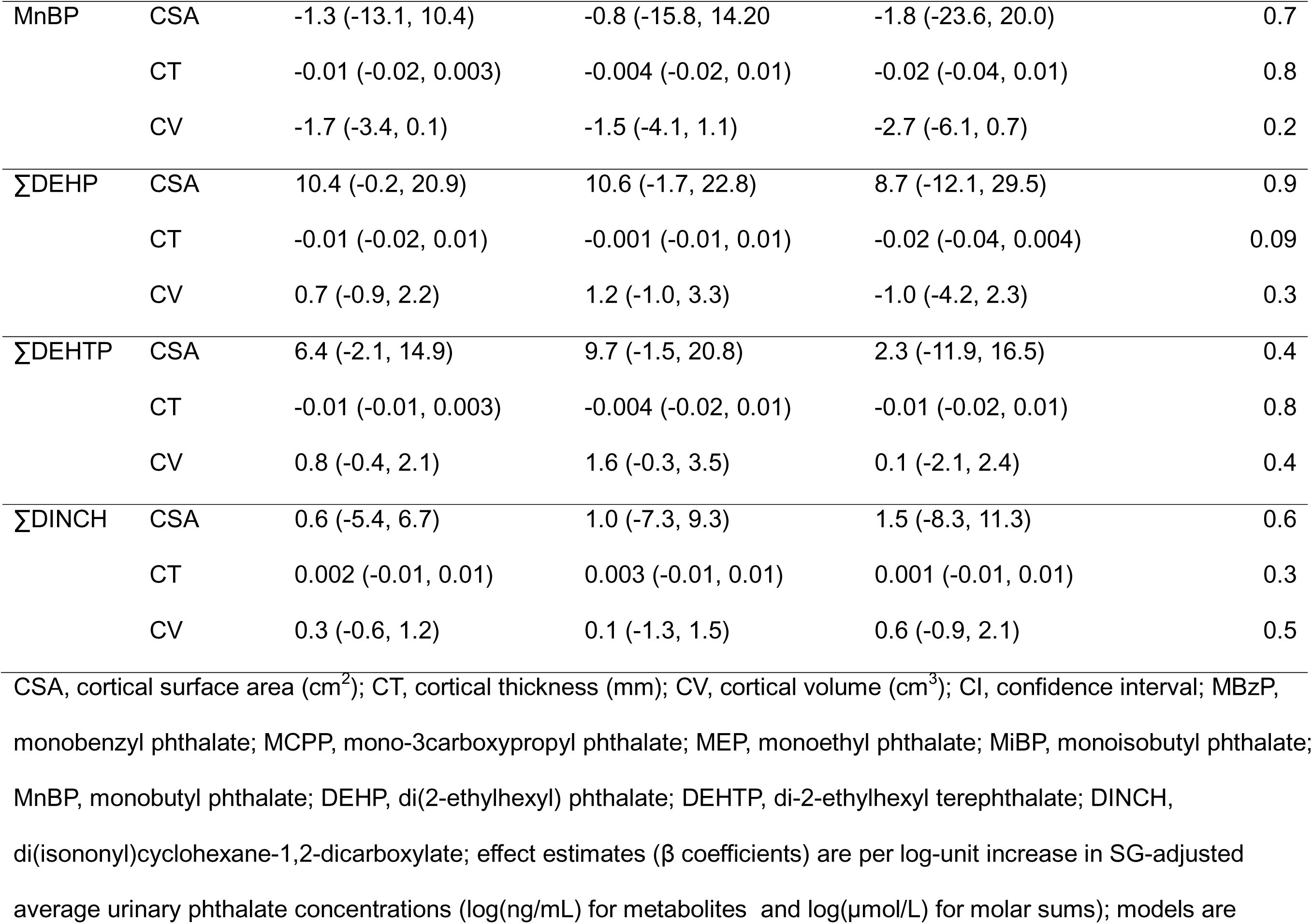
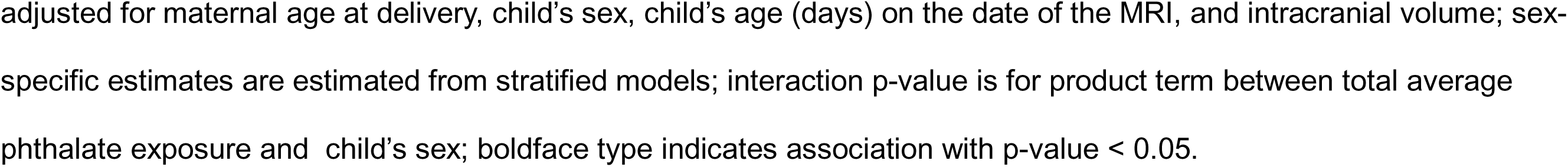
Associations of total average phthalate exposures with cortical surface area, thickness, and volume in the UNC BCP (n=369).

When considering sex-specific associations, main effects appeared to be driven by males (Figure 1). Among males, we observed inverse associations with GMV (MEP: −3.0 cm^3^, 95% CI: −5.9, −0.1; MiBP: −2.4 cm^3^, 95% CI: −4.6, −0.2) and a positive association between ∑DEHP and WMV (4.5 cm^3^, 95% CI: 0.1, 8.9) (Table 2). Although there was no main effect between ∑DINCH and WMV in the overall sample, among females ∑DINCH was associated with a 2.3 cm^3^ increase in WMV (95% CI: 0.3, 4.2, interaction p-value=0.001). We also observed heterogeneity by sex in the relationship between ∑DEHP and GMV (interaction p-value=0.03), with a suggestive positive association among females (2.2 cm^3^, 95% CI: −0.05, 4.5) and a nonsignificant inverse association among males (−2.5 cm^3^, 95% CI: −6.3, 1.2). There were no statistically significant sex differences for the cortical measures (Table 3), though differences were suggestive for MBzP with CSA (interaction p-value=0.07) and ∑DEHP with CT (interaction p-value=0.09). Further, the main effect with MiBP and lower cortical thickness was also observed when restricted to males only (−0.02 mm, 95% CI: −0.03, −0.004).

In sensitivity analyses of cumulative average exposure, patterns of associations were similar to those reported in main analyses using total average exposure, albeit most were slightly attenuated (Supplemental Tables 1-2, Supplemental Figure 2). When we attempted to assess critical windows of exposure by dichotomizing the sample at ages 12- and 18-months, analyses were underpowered and models encountered convergence problems. As such, we were unable to interpret these analyses (data not shown). In age-stratified analyses dichotomized at two years old (Supplemental Table 3), we did not observe evidence of a statistical interaction between phthalate/replacement plasticizer exposure and age (divided at two years). Main effects that we observed with GMV in the overall sample were observed in the younger age group only (MCPP: −2.7 cm^3^, 95% CI: −4.7, −0.6; MiBP: −2.3 cm^3^, 95% CI: −4.1, −0.4). Conversely, the positive association between DEHP and WMV in the main sample appeared to be driven by children older than two years (3.9 cm^3^, 95% CI: 0.5, 7.4). When we corrected p-values for multiple testing, no associations passed the FDR threshold (data not shown).

## Discussion

We found that higher early life urinary concentration of MCPP was associated with lower GMV, and ∑DEHP was associated with higher WMV, in children two weeks to seven years old. Relationships between phthalates and global brain morphology were more apparent among males. There was a general pattern of inverse associations between phthalate metabolite concentrations and GMV among males (e.g., MCPP, MEP, MiBP, and MnBP), though some associations were not statistically significant. Notably, the replacement plasticizer DINCH was associated with higher WMV among females only. With the exception of MiBP and slightly lower cortical thickness, we did not observe effects of phthalate/plasticizer exposure on cortical measures. In age-stratified analyses, we observed associations with lower GMV (e.g., MCPP and MiBP) in children under age two, and associations between DEHP and higher WMV in children older than age two. No associations remained significant when we corrected p-values for multiple hypothesis testing (n=40 tests). Because selection of exposures and outcomes was hypothesis-driven, we prioritized results that were not FDR-adjusted.

The present study is the first to examine early life phthalate exposures in association with brain MRI in a longitudinal setting. Strengths of our approach include repeated biomarkers of exposure across the study period, repeated neuroimaging with dense longitudinal sampling during sensitive periods of development, sufficient sample size to evaluate sexually dimorphic associations, and inclusion of the emerging replacement plasticizers DINCH and DEHTP. We note that our study also has important limitations. We elected to use average phthalate concentrations to better represent typical exposure across the study period because phthalates are non-persistent chemicals with short half-lives. Intraclass correlations for phthalate metabolites and replacement plasticizers among participants with multiple samples in the UNC BCP were previously reported (range, ρ = 0.10–0.48), indicating low to moderate reliability and high within-individual variation^18^. In sensitivity analyses, we evaluated phthalates as average cumulative exposures until each MRI scan date to determine whether exposure information obtained after the index MRI was influencing results. Interpretation of these analyses were similar to those estimated using total average exposures, suggesting that exposure measurement error did not influence results. To maximize power, we presented total average exposures as primary analyses (n_total_=369 versus n_cumulative_=284), with cumulative average exposures as supplemental information. While age-specific analyses were underpowered, previous analyses of phthalate and plasticizer exposure levels in this population revealed that concentrations of MnBP, MCPP, MiBP, MEP, and MBzP were inversely associated with age, and often highest before age one^18^. Based on this pattern, and the age-dependence of brain growth during early life, we hypothesized that critical windows may exist in the relationship between phthalates and brain morphology. However, the temporal distribution of data undermined our ability to evaluate exposure-age interactions because age-stratified sample sizes were prohibitively small. Finally, owing to study protocol constraints, most UNC BCP participants lived in close proximity to UNC and the study population over-represents white participants of higher socioeconomic and educational backgrounds. This lack of diversity limits the generalizability of our results to more diverse populations, particularly in the context of racial/ethnic and socioeconomic disparities in phthalate exposure^42^ and socioeconomic impacts on early life brain growth^43,44^. It is possible that associations may have been more stark in populations with higher exposure and fewer community and family resources capable of mitigating harms from environmental toxicants.

Although no other studies have assessed postnatal phthalate exposure with neuroimaging at the early ages we evaluated, one study evaluated prenatal maternal urinary phthalates in association with global brain volumes at age 10 in the Generation R cohort (n=775)^27^. The authors reported inverse relationships between prenatal concentrations of MiBP (β =−8.11, 95% CI: −13.56, −2.66) and MEP (β =−10.7, 95%CI: −18.12, −3.28) with GMV. In our analysis, we also observed inverse associations for these metabolites and GMV, however these associations were observed among males only in our sample. Interestingly, sex-specific effects were observed predominantly among females in Generation R, whereas sex-specific patterns were more apparent among males in our analyses. An imperfect comparison, this discrepancy may be related to sex differences in timing of exposure (prenatal versus postnatal) or neurodevelopment (infancy and childhood versus adolescence). Because the Generation R study didn’t measure DINCH, we couldn’t compare our observed association with higher WMV in females.

Many studies linking phthalate exposures with altered neurodevelopment, including externalizing behaviors and attention-deficit hyperactivity disorder (ADHD), find that boys are often at higher risk for poorer neurodevelopmental outcomes^22^. Though the endocrine-disrupting properties of phthalates are well known^45^, modification of associations with neurodevelopment by sex is variable across studies. These inconsistencies may be due to heterogeneity in neurodevelopmental endpoints and assessments. In the present study, we observed sex-specific effects, linking multiple phthalate metabolite exposures (e.g., MEP, MiBP, and DEHP) to reduced GMV among boys. Taken together with prior work reporting associations between prenatal MiBP and adverse executive function and behavioral problems in preschool-aged^46^ and 6-10 year old boys^47^, these results suggest that sex-specific alterations in gray matter growth could possibly underlie relationships between MiBP and neurobehavior.

A robust literature links regional and global gray matter deficits to inattention, hyperactivity, impulsivity, and ADHD. Vetter et al. report lower GMV in boys with ADHD and oppositional defiance disorder compared to typically developing boys aged 11-17 years old^48^. A study in the Adolescent Brain Cognitive Development Study (n= 10,692) found that inattentive, hyperactivity, and impulsivity traits show GMV deficits in areas associated with attention and emotion regulation, visuospatial processing and memory, and motor activity at 9-10 years of age^49^. A 2024 meta-analysis found that pediatric bipolar disorder and ADHD share decreased regional GMV in areas related to emotion processing and attention^50^. We offer evidence that the neural substrates connecting phthalate exposures and neurobehavioral outcomes could involve disruptions in gray matter development. Moreover, endocrine disrupting properties seem to be eliciting stronger effects on these neural substrates, and their subsequent neurobehavioral endpoints, in boys.

The neurotoxicity of DEHP has been studied extensively, with frequently reported sex differences in associations with neurobehavior and autism spectrum disorders^51^. We observed that DEHP concentrations were associated with higher WMV overall, and lower GMV among males. In the APrON study (n=448), higher exposure to maternal prenatal DEHP was associated with poorer motor and executive function at two years old, with worse outcomes in boys^52^. A subset of this study (n=76) acquired neuroimages at 3-5 years of age and found that phthalate exposures were associated with impaired white matter microstructure integrity^26^. In a study of six-year-olds in Korea, childhood DEHP exposure had an adverse effect on IQ and attentional performance (n=175)^53^. A cross-sectional analysis of phthalates and MRI at ages 6-15 (n=115) found that DEHP metabolites were: negatively correlated with cortical thickness in some regions, significantly higher in boys with ADHD than in boys without ADHD, and the metabolite concentration was positively correlated with the severity of externalizing symptoms^29^. Despite being regulated in childhood toys and childcare products, we observed that children continue to be exposed to DEHP, likely due to diet being a major source of exposure^54^. Moreover, this early life exposure to DEHP impacts structural brain growth with potential implications for future neurodevelopment across early childhood and into adolescence.

In summary, our findings demonstrate that early life phthalate exposure impacts structural brain growth in the first years of life, particularly among males. The associations and sex differences we note here contribute to understanding the neural basis through which phthalates disrupt emotional, behavioral, and cognitive development. The observed association between DINCH and WMV suggests that the safety of replacement plasticizers should be carefully considered. Future studies should assess regional and subcortical measures, functional connectivity, and neurobehavioral outcomes.

## Data Availability

All data produced in the present study are available upon reasonable request to the authors

## Acknowledgement of funding

This research was supported in part by the Intramural Research Program of the NIH, EPA (RD-84021901), NIH/NIEHS (R01 ES033518, P30 ES010126, K99 ES035123), and a University of North Carolina, Gillings School of Global Public Health “Gillings Innovation Lab” award. The Baby Connectome Project was funded by U01 MH110274.

## Supplemental Tables & Figures

**Supplemental Figure 1.**
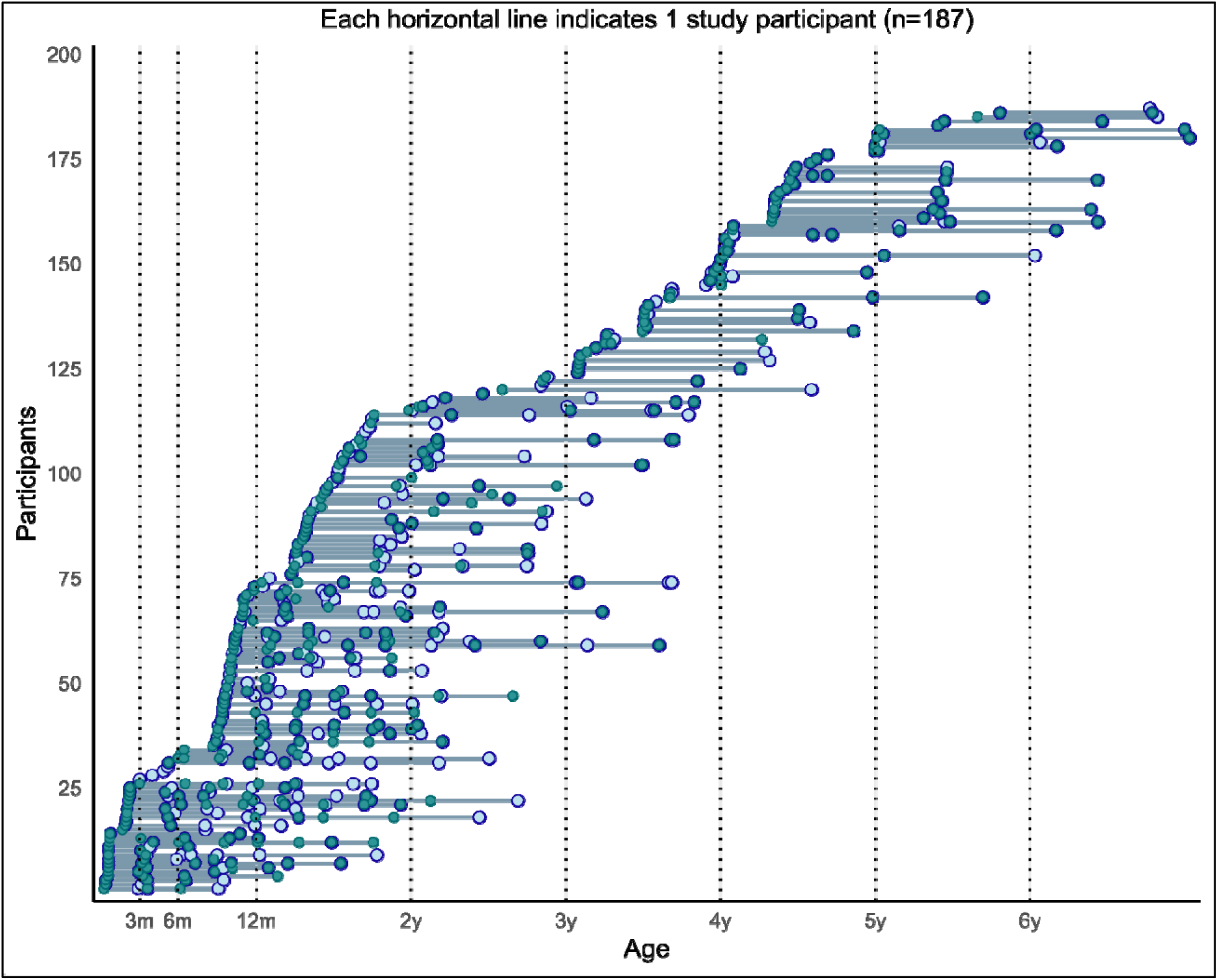
Enrollment in the UNC BCP from 2017-2020. Blue circles represent urine samples, green circles represent MRIs, and green circles with blue outlines represent simultaneous urines and scans.

**Supplemental Figure 2.**
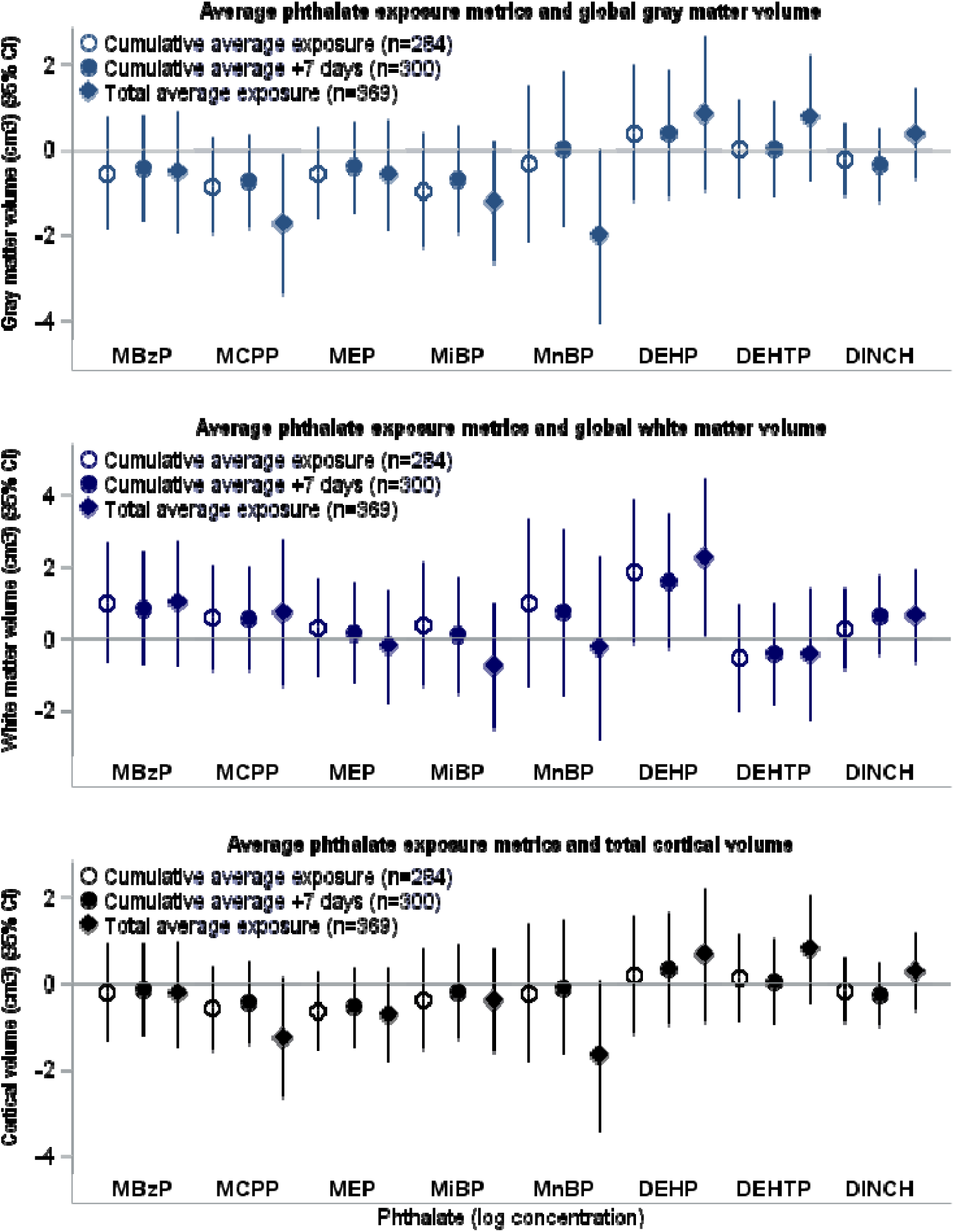
Associations between different average phthalate exposure metrics (cumulative average, cumulative average plus seven days after scan, and total average across th study period) and global brain volumes (gray matter, white matter, total cortical).

**Supplemental Table 1.**
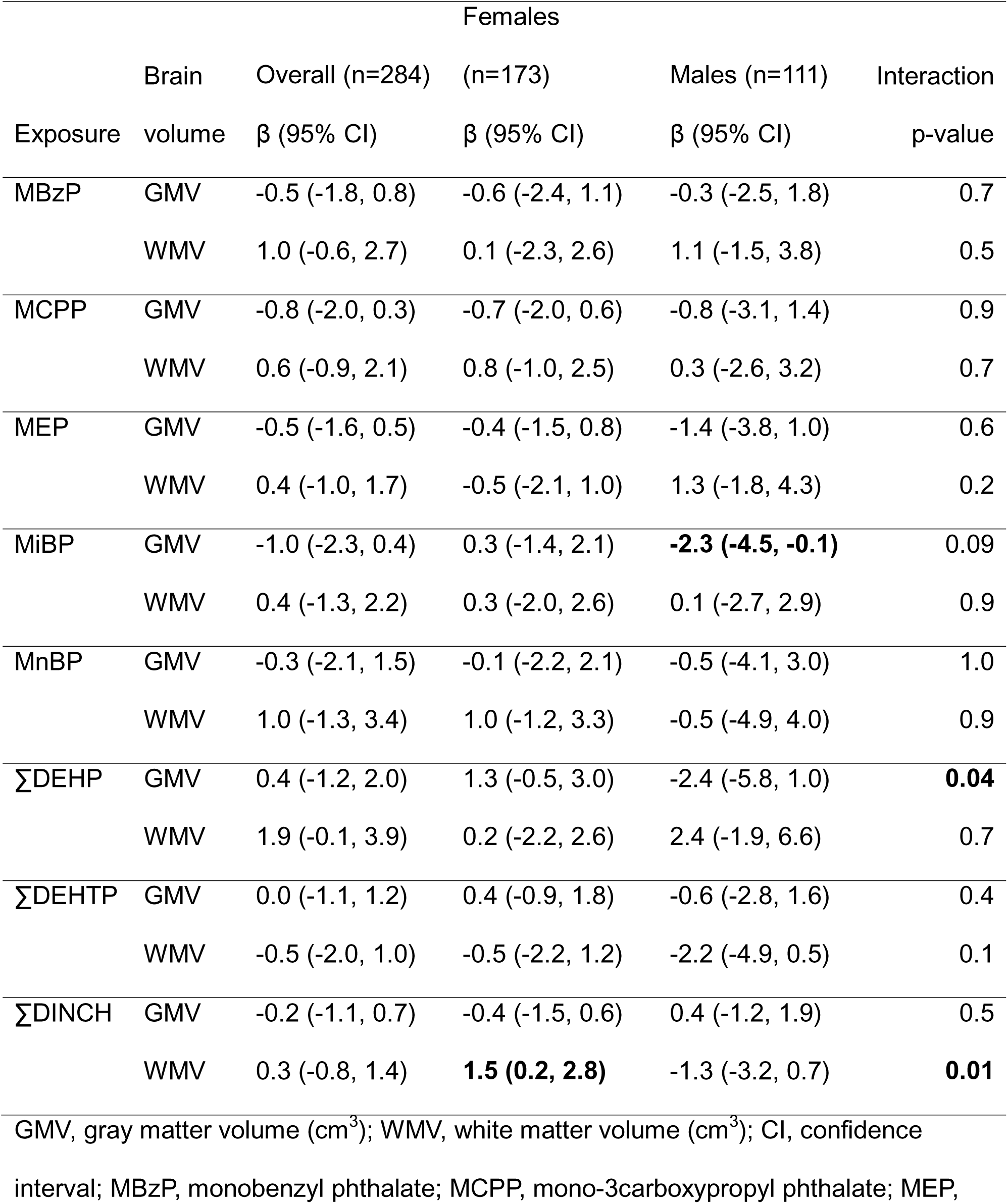
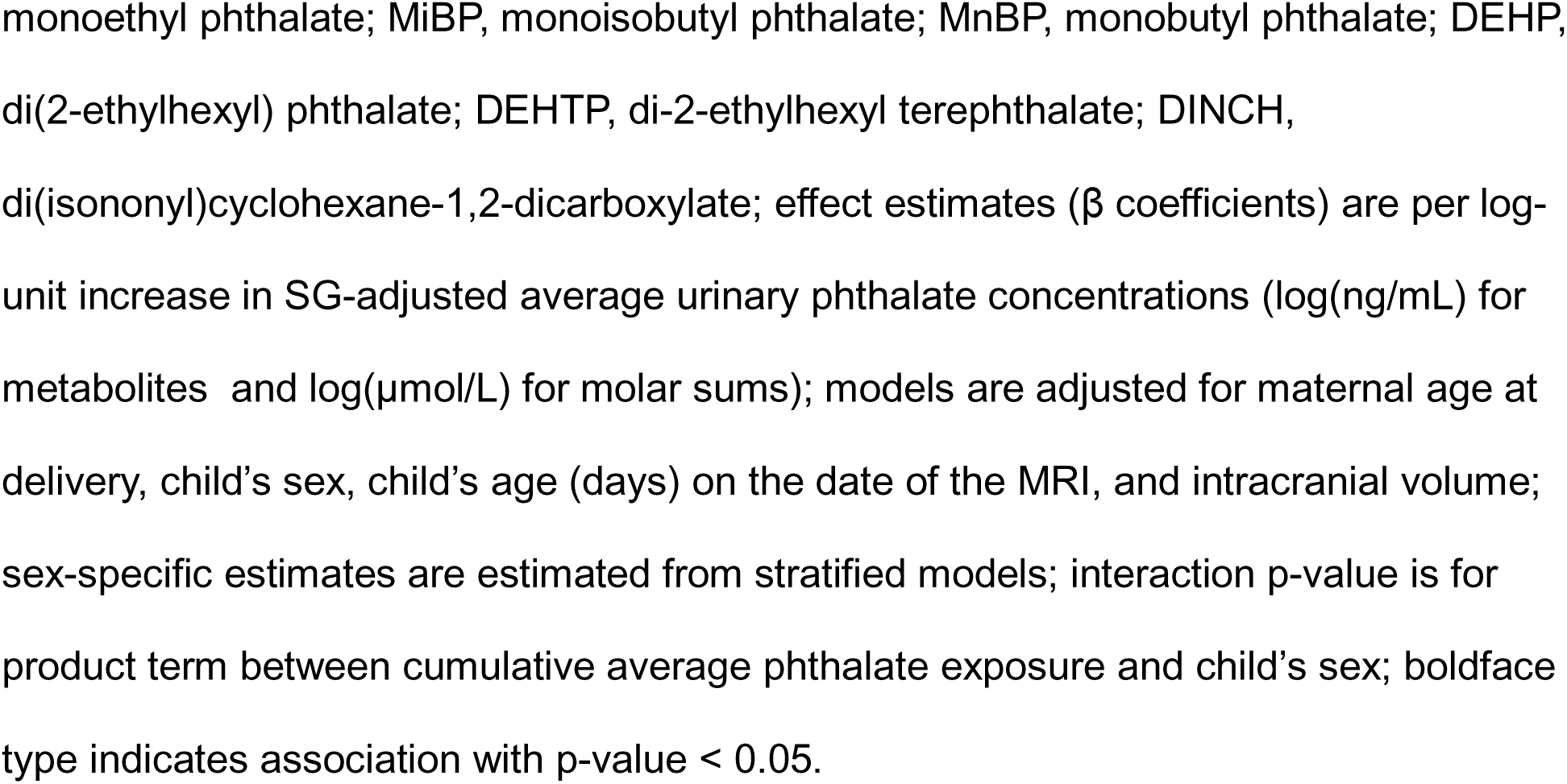
Associations of cumulative average phthalate exposures with global gray and white matter volumes in the UNC BCP (n=284).

**Supplemental Table 2.**
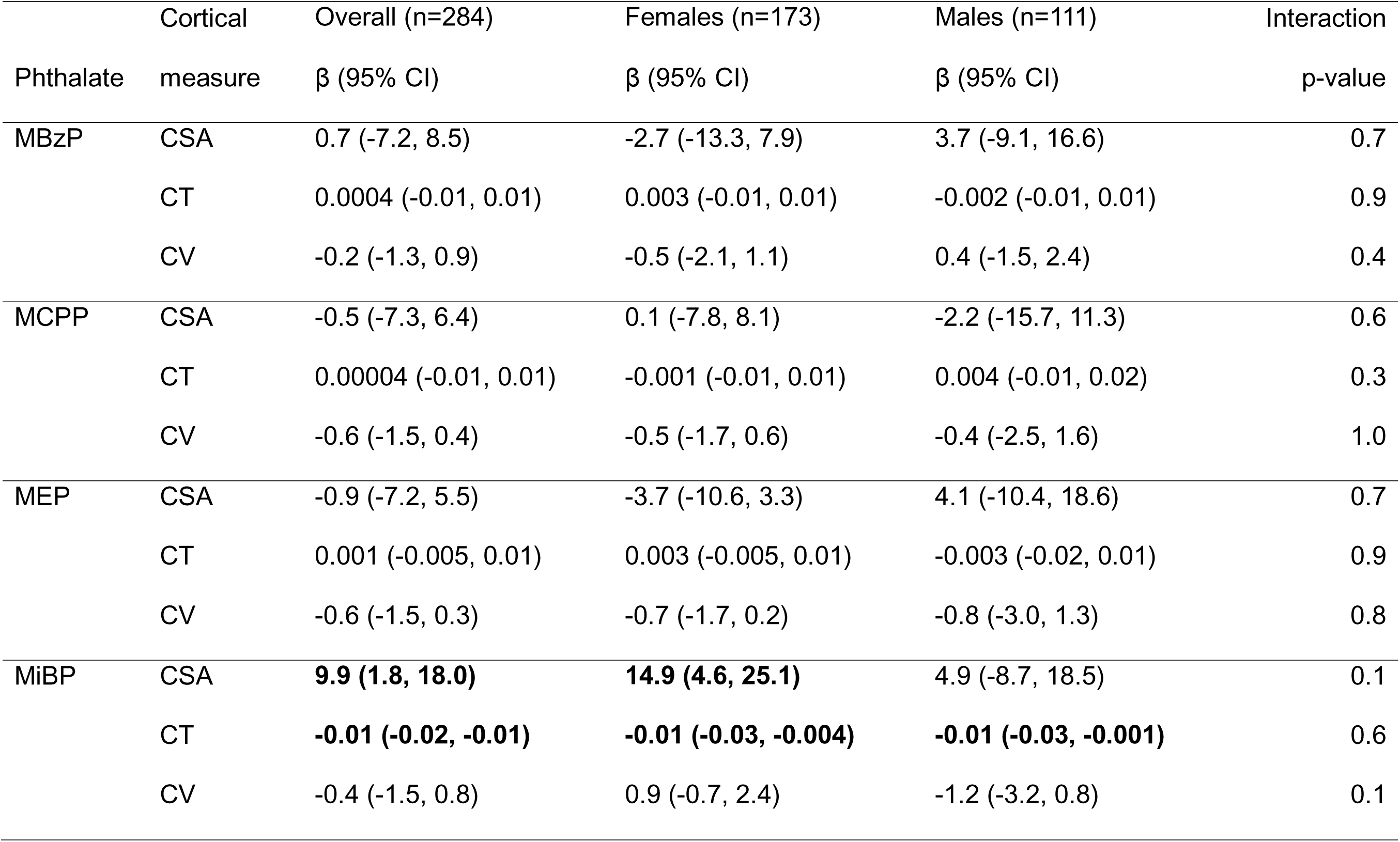
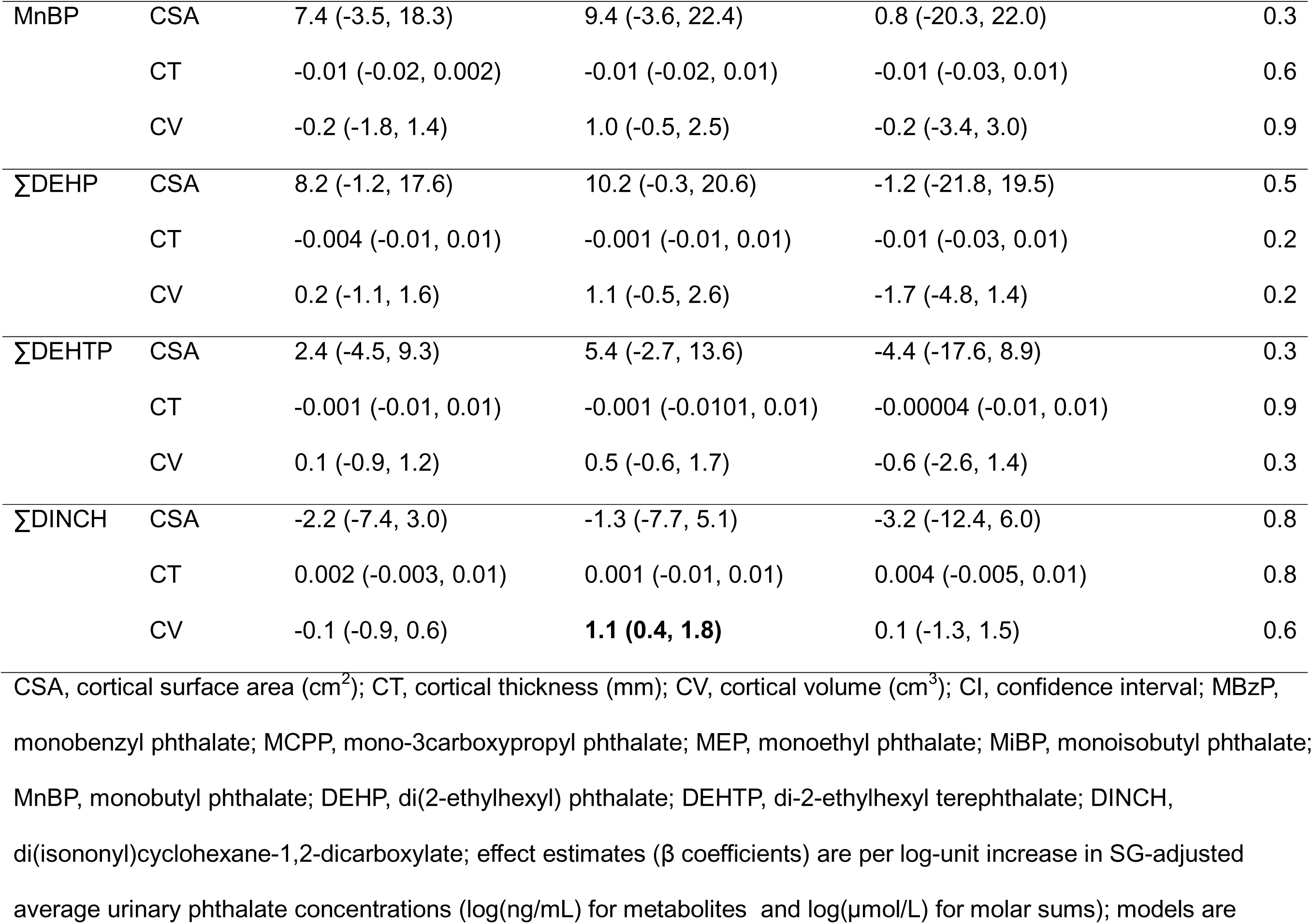
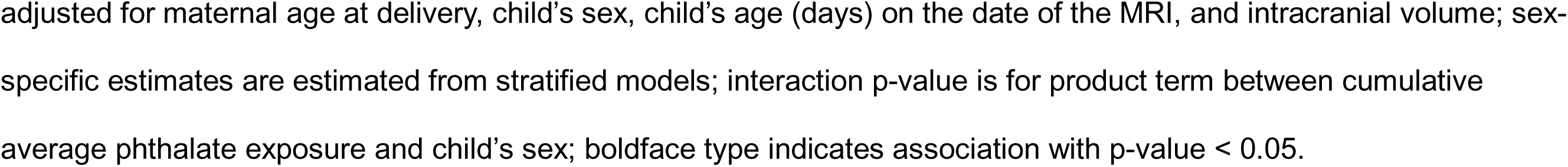
Associations of cumulative average phthalate exposures with cortical surface area, thickness, and volume in the UNC BCP (n=284).

**Supplemental Table 3.**
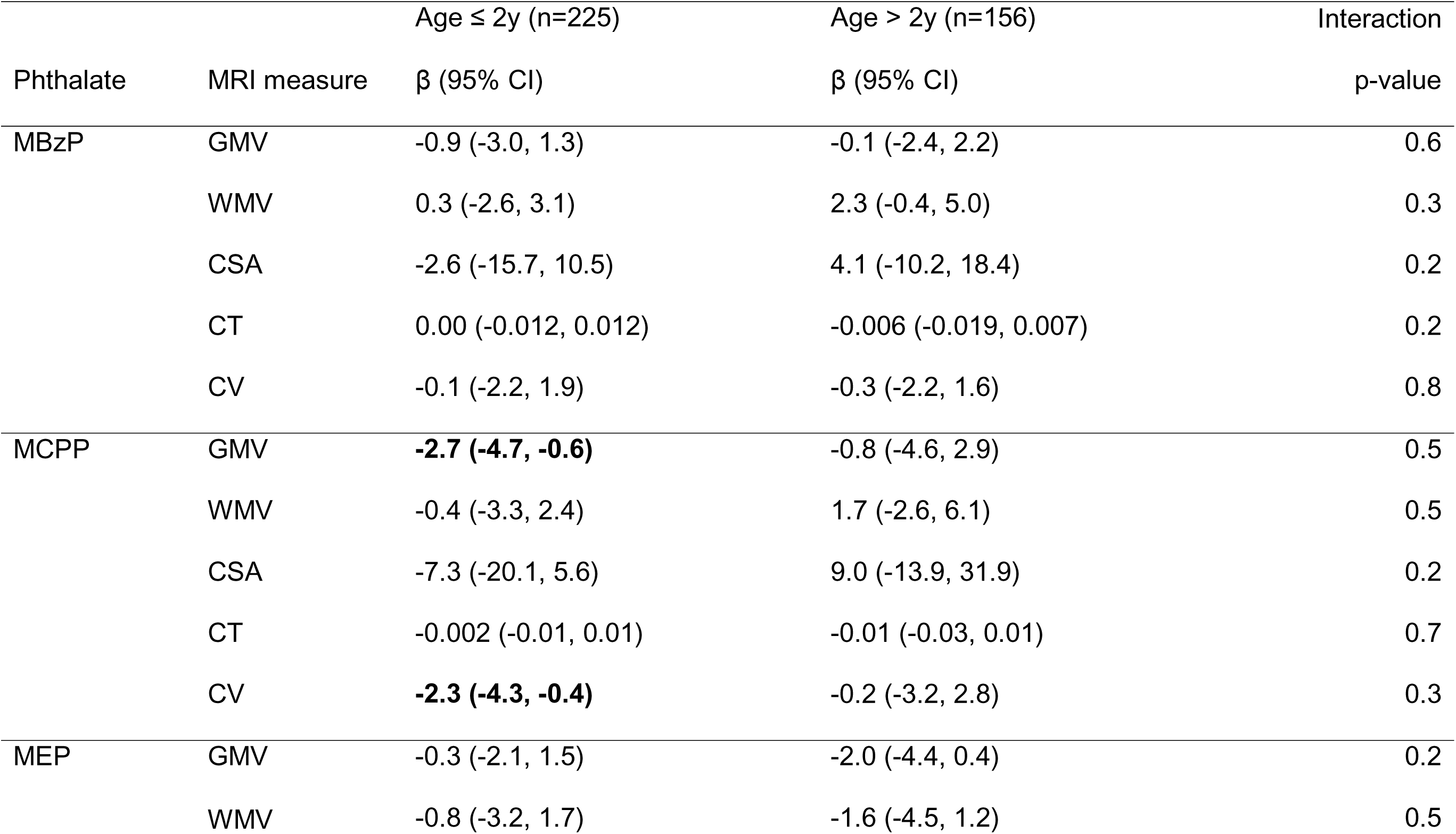
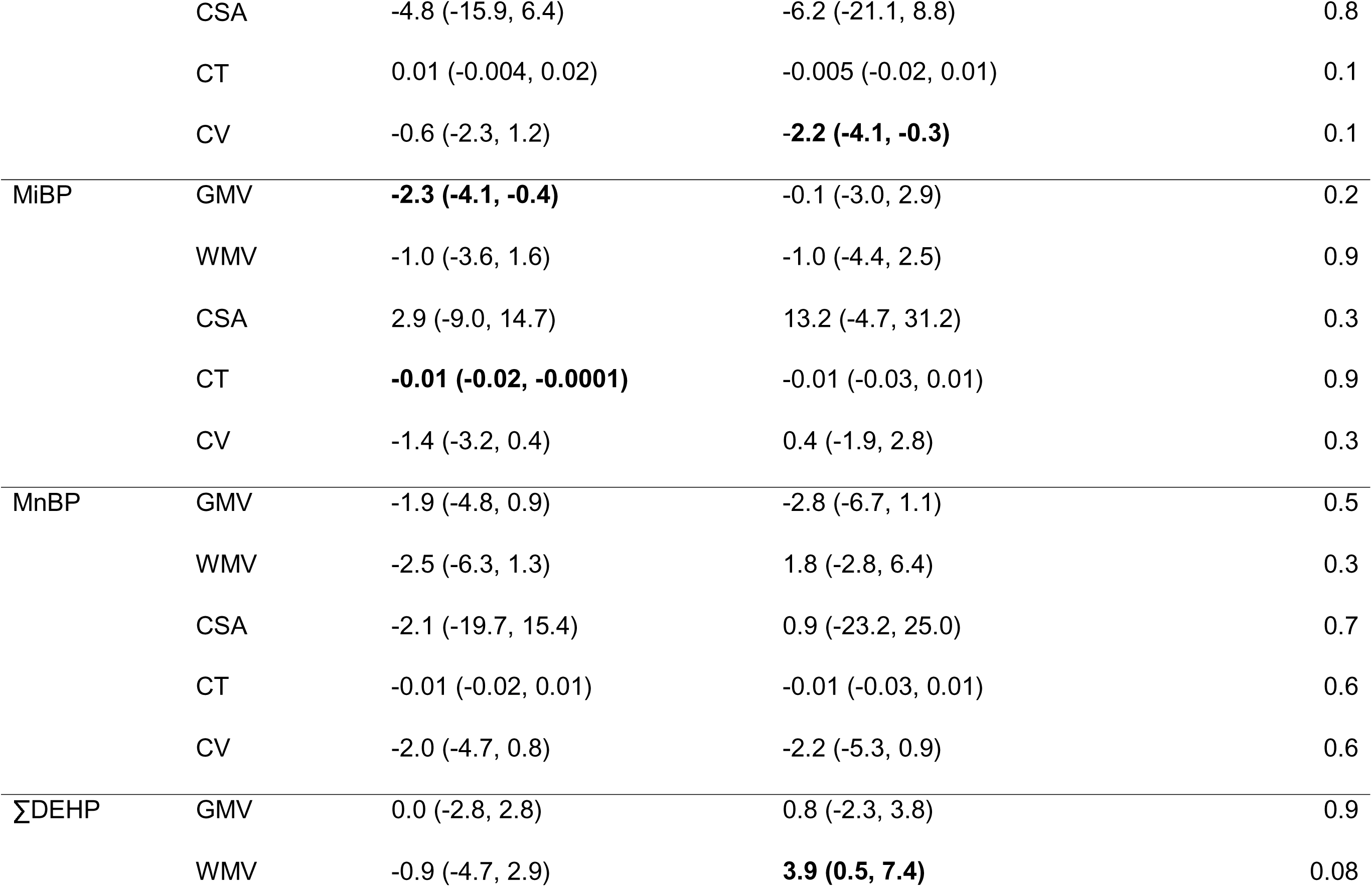
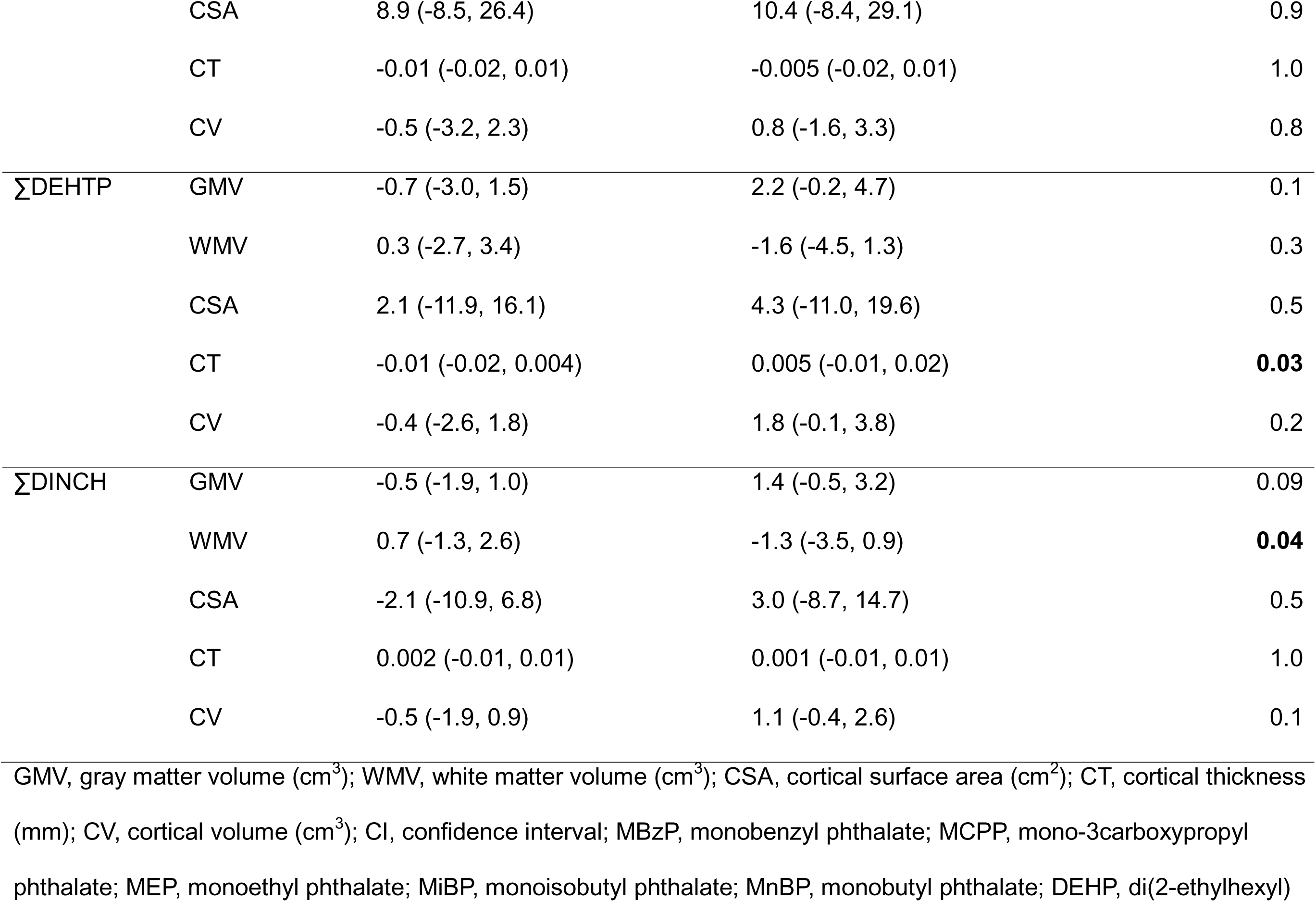
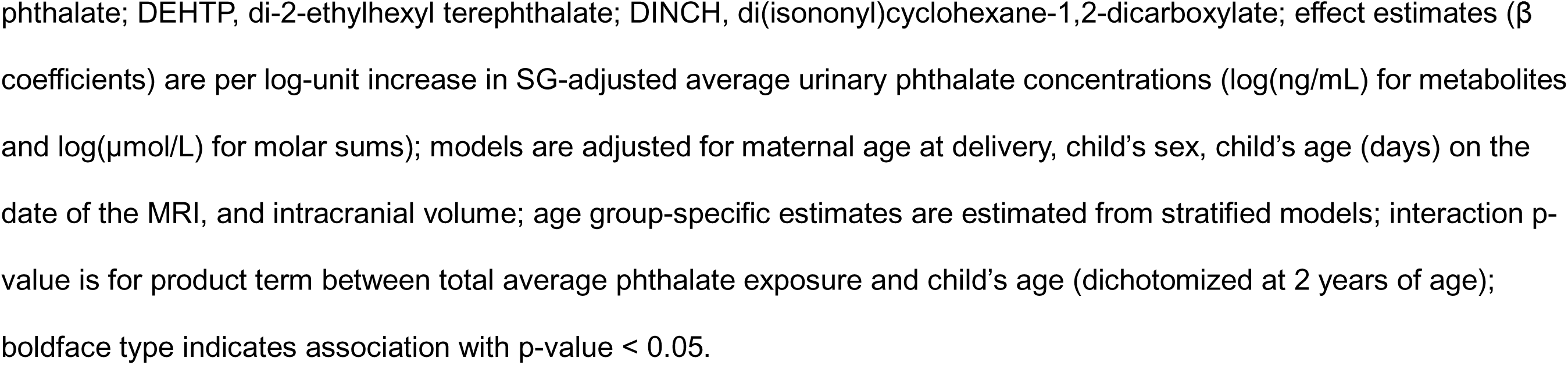
Age-stratified associations of total average phthalate exposures with gray and white matter volume, and cortical surface area, thickness, and volume dichotomized at age 2 years old among children in the UNC BCP (n=225).

## Notes

### Competing Interest Statement

The authors have declared no competing interest.

### Author Declarations

Ethics committee/IRB of the University of North Carolina at Chapel Hill gave ethical approval for this work

